# Identifying weather variables as important clinical predictors of bacterial diarrhea among international travelers to Nepal and Thailand

**DOI:** 10.1101/2021.12.16.21267958

**Authors:** Melissa A. Pender, Timothy Smith, Ben J. Brintz, Prativa Pandey, Sanjaya Shrestha, Sinn Anuras, Samandra Demons, Siriporn Sornsakrin, James A. Platts-Mills, Ladaporn Bodhidatta, Daniel T. Leung

## Abstract

**Background:** Clinicians and travelers often have limited tools to differentiate bacterial from non-bacterial causes of travelers’ diarrhea (TD). Development of a clinical prediction rule assessing the etiology of TD may help identify episodes of bacterial diarrhea and limit inappropriate antibiotic use. We aimed to identify predictors of bacterial diarrhea among clinical, demographic, and weather variables, as well as to develop and cross-validate a parsimonious predictive model.

**Methods:** We collected de-identified clinical data from 457 international travelers with acute diarrhea presenting to two healthcare centers in Nepal and Thailand. We used conventional microbiologic and multiplex molecular methods to identify diarrheal etiology from stool samples. We used random forest and logistic regression to determine predictors of bacterial diarrhea.

**Results:** We identified 195 cases of bacterial etiology, 63 viral, 125 mixed pathogens, 6 protozoal/parasite, and 68 cases without a detected pathogen. Random forest regression indicated that the strongest predictors of bacterial over viral or non-detected etiologies were average location-specific environmental temperature and RBC on stool microscopy. In 5-fold cross-validation, the parsimonious model with the highest discriminative performance had an AUC of 0.73 using 3 variables with calibration intercept -0.01 (SD 0.31) and slope 0.95 (SD 0.36).

**Conclusions:** We identified environmental temperature, a location-specific parameter, as an important predictor of bacterial TD, among traditional patient-specific parameters predictive of etiology. Future work includes further validation and the development of a clinical decision-support tool to inform appropriate use of antibiotics in TD.

## Introduction

Travelers’ diarrhea (TD) is a common illness affecting approximately 20% of international travelers and contributing to one third of post-travel medical visits.^1^ Etiologies include bacterial, viral, parasitic, or mixed pathogens and can be influenced by external factors such as season,^2^ location, or activity. In the context of international travel, laboratory diagnosis of TD may be costly, inconvenient, or unavailable. Current joint guidelines from the International Society of Travel Medicine and the US Centers for Disease Control recommend providing travelers with standby antibiotics for self-treatment in the event of diarrheal symptoms.^3^ Such treatment is currently guided by severity of illness based on functional impact, with limited consideration of etiology, resulting in potential for inappropriate use of antibiotics for severe viral or protozoal diarrheal infections.^4^ Inappropriate use of antibiotics can lead to side effects, development of resistance,^5^ and the potential for other infections including *C. difficile*.^6^ Clinical decision support tools incorporating clinical prediction rules provide an attractive alternative to microbial testing for identification of cases of bacterial TD when laboratory availability or access is limited.

A clinical prediction rule (CPR) is a tool predicting the probability of a specific outcome. Recent advances in machine learning methods such as random forests offer new tools for developing clinical prediction models.^7-9^ Prior studies using CPRs to predict bacterial diarrhea among children,^8, 10-12^ and travelers,^13^ have demonstrated promising results but have been limited by small sample sizes, sub-optimal performance characteristics of predictors, and limited pathogen identification. Development of new diagnostic tools such as the TaqMan® array card have recently improved identification of diarrheal pathogens.^14^ In this study, we incorporated machine learning methods with traditional statistical methods to develop and cross-validate a predictive model with the ultimate goal of developing a parsimonious CPR to assess the etiology of TD from primarily clinical and epidemiological parameters.

## Methods

### Ethical approval

The institutional review boards of the University of Utah, the Walter Reed Army Institute of Research, CIWEC Hospital, the Nepal Health Research Council (NHRC), and Bumrungrad International Hospital, approved this study.

### Study subjects

We collected de-identified data from studies of international travelers seeking care for acute diarrhea at two single-hospital sites located in Nepal and Thailand. The Bumrungrad International Hospital in Bangkok, Thailand enrolled 173 travelers from Feb. 2012 – Dec. 2014 as part of a study comparing TD diagnostic tests.^14^ The CIWEC Travel Medicine Center in Kathmandu, Nepal enrolled 284 travelers from March 2016 - May 2017.

Among both populations, inclusion criteria consisted of adult travelers ≥ 18 years old who were citizens of countries in North America, Europe, Australia, New Zealand, Japan, Taiwan, or South Korea and who presented seeking healthcare for acute diarrhea or gastroenteritis.^14^ We included all subjects who met definitions of acute diarrhea or acute gastroenteritis. Acute diarrhea was defined as ≥3 loose/liquid stools in the preceding 24 hours OR ≥2 loose/liquid stools in the preceding 24 hours PLUS at least ≥2 associated GI symptoms, including subjective fever/chills, nausea, vomiting, abdominal cramping, abdominal pain, tenesmus, bloating, fecal urgency or gross blood in stool. Acute gastroenteritis was defined as ≥3 loose/liquid stools in the preceding 24 hours with ≥1 additional GI symptoms as above OR ≥3 vomiting episodes in the preceding 24 hours with ≥1 additional GI symptoms OR Vomiting (≥2) OR diarrhea (≥2) within the preceding 24 hours with ≥2 additional GI symptoms.^1, 3^ Travelers were excluded if they were not citizens of the countries listed above or if they had resided outside of their country of citizenship for greater than 1 year. Additional exclusion criteria included symptoms of diarrhea present for greater than 7 days, an inability to provide a stool sample, or an inability to provide written informed consent.^14^

### Data collection

After obtaining written informed consent, travelers provided a stool sample and completed a questionnaire covering demographics, clinical history, and associated symptoms (Table 1). Stool samples were sent to the laboratory for stool microscopy as well as additional diagnostic testing detailed below. Stool microscopy variables included the presence or absence of red blood cells, white blood cells, and/or stool mucus.

**Table 1.** Demographics, symptoms, and stool microscopy results collected from travelers to Nepal and Thailand.

| Variable | Nepal (% of n=284) | Thailand (% of n=169) |
| --- | --- | --- |
| Male | 123 (43) | 106 (62) |
| Median age (IQR) | 26 (10) | 32 (5) |
| North American nationality | 78 (27) | 32 (19) |
| European nationality | 158 (56) | 71 (42) |
| Australia/New Zealand nationality | 38 (13) | 26 (15) |
| Asian nationality | 10 (3.5) | 40 (24) |
| Median site temperature °F (30-day average) (IQR) | 70.3 (12) | 84.2 (3.2) |
| Median site precipitation (30-day average) in inches (IQR) | 0 (0.09) | 0 (0.14) |
| Diarrhea present | 280 (99) | 169 (100) |
| Median diarrhea duration in hours (IQR) | 48 (59) | 24 (52) |
| Median total # diarrhea (IQR) | 17 (17) | 8 (5) |
| Vomiting | 165 (58) | 87 (51) |
| Abdominal cramping/pain | 244 (86) | 145 (86) |
| Malaise/Fatigue | 267 (94) | 149 (88) |
| Febrile | 40 (14) | 103 (61) |
| Bloody Stool | 12 (4.2) | 19 (11) |
| Prior treatment | 87 (31) | 91 (54) |
| Stool grade: formed | 2 (0.7) | 0 (0) |
| Stool grade: soft | 14 (4.9) | 12 (7.1) |
| Stool grade: loose | 62 (22) | 81 (48) |
| Stool grade: watery | 206 (73) | 76 (45) |
| Stool microscopy: Mucus | 215 (76) | 73 (43) |
| Stool microscopy: WBC | 227 (80) | 135 (80) |
| Stool microscopy: RBC | 52 (18) | 56 (33) |
| Stool microscopy: Parasite | 45 (16) | 1 (0.6) |

To identify diarrheal pathogens, we relied on multiple diagnostic techniques including stool culture, real-time polymerase chain reaction (PCR), and enzyme-linked immunosorbent assay (ELISA).^14^ Using standard laboratory culture techniques, stool culture was used to isolate *Aeromonas, Arcobacter, Campylobacter, Escherichia coli (E. coli), Plesiomonas, Salmonella, Shigella*, or *Vibrio* species (Table 3). *E. coli* strains were identified based on stool culture followed by PCR detecting targets for enterotoxigenic (ETEC), enteropathogenic (EPEC), shiga toxin-producing (STEC), or enteroaggregative (EAEC). Reverse transcription PCR identified norovirus and sapovirus. We identified parasitic infections such as *Cryptosporidium, Entamoeba histolytica*, and *Giardia* using ELISA kits.

In addition to conventional diagnostic methods, data collected from Bumrungrad International Hospital in Thailand included organisms detected on an enteric pathogen panel TaqMan® array card (TAC). TAC testing detects multiple enteropathogens using real-time PCR.^15, 16^ We used the QiaAmp Fast Stool DNA kit to extract total nucleic acid and used reagents from the Ag-Path-ID One-Step RT-PCR kit. Our TAC panel detects the following 12 species of bacteria: *Aeromonas, Bacteroides fragilis, Campylobacter coli*/*C. jejuni, Clostridium difficile*, enteroaggregative *Escherichia coli*, enteropathogenic *E. coli*, enterotoxigenic *E. coli, Helicobacter pylori, Salmonella, Shigella*/enteroinvasive *E. coli*, Shiga toxin–producing *E. coli*, and *Vibrio cholerae*. It also detects the following five viruses (adenovirus, astrovirus, norovirus GI/GII, rotavirus, and sapovirus), five species of nematodes (*Ancylostoma duodenale, Ascaris lumbricoides, Necator americanus, Strongyloides stercoralis*, and *Trichuris trichiuria*), five protozoan parasites (*Cryptosporidium, Cyclospora cayetanensis, E. histolytica, Giardia lamblia*, and *Cystoisospora* belli), and two species of fungi (*Encephalitozoon intestinalis* and *Enterocytozoon bieneusi*).^14^ We identified pathogen positivity using a positive threshold cycle (Ct) value cutoff of <35; a negative result consistent of a Ct value >35. Further details regarding TAC analysis and comparison to conventional methods were described in prior reports.^14, 17^

In order to investigate the effect of seasonality and local weather patterns on TD etiology, we constructed variables of temperature and precipitation. We obtained date-specific site temperature and precipitation data from National Oceanic and Atmospheric Administration (NOAA) sites in Bangkok and Kathmandu. To create a temperature variable, we averaged the temperature for the 14 days prior to patient enrollment. The same process was used for the precipitation variable.

### Statistical analysis and modeling

In the construction of our models, we used binary variables with the exception of continuous variables age, diarrhea duration, number of diarrheal episodes, average temperature, and average precipitation. Variables with >40% missingness were removed and multiple imputation was used to account for any additional missing data. Four patient cases from the Nepal dataset were excluded from analysis due to >40% data missingness leaving a total of 453 cases for analysis. For each case, we categorized the etiology of diarrhea as bacterial-only infection, mixed infection with both bacterial and non-bacterial organisms, viral-only infection, protozoal/parasite-only infection, or infection with no identified organism.

All analyses were completed using R,^18^ and model development/validation was completed in accordance to the TRIPOD checklist (supplemental table S1).^19^ Using the combined dataset from both sites, we used random forest regression consisting of 1000 decision trees and the default number of variables considered at each split (p/3, where p is the number of predictors considered) to determine variable importance. This method uses multiple decision trees to determine and calculate the reduction in mean squared prediction error for each variable. We arranged the variables in order of descending importance according to the reduction in mean squared prediction error. As a secondary analysis, to examine the top predictors of each site, given the smaller sample sizes, we used univariable methods (Chi-square or Fisher’s exact tests for categorical variables and Wilcoxon rank-sum tests for continuous variables) to determine the relationship between variables and bacterial-only cases at each site.

To assess predictive performance, we used 5-fold repeated cross-validation with 100 iterations using both logistic regression and random forest models. In each iteration, we randomly divided data combined from all sites into an 80% training set and a 20% testing set. We calculated the area under the receiver operator curve (AUROC or AUC) for both the logistic regression (LR) and the random forest (RF) models using a range of variables from 1 to 35 in varying increments. We fit models with varying input parameter sets and outcomes to determine the best discriminative performance based on the AUC. For each cross-validation test set, we estimated the calibration intercept and slope, respectively, by first fitting a logistic regression with the predicted value of each test observation as an offset in an intercept-only model, and second by fitting a logistic regression model with the predicted value of each test observation as the regressor.

## Results

We collected demographic, symptom, and stool microscopy data from the single-hospital site in Thailand from Feb 2012 to Dec 2014 and from the single-hospital site in Nepal from March 2016 to May 2017 (Table 1). Among the 457 cases, we detected the following diarrheal pathogens; 195 bacterial cases, 125 cases with mixed pathogens, 68 cases without detected pathogen, 63 viral cases, and 6 cases of protozoal disease (Table 2). These etiologies are further broken down into number per month of data collection in Thailand and Nepal (Figures S1 and S2, respectively), and compared with 30-day moving averages of environmental temperature and precipitation (Figures S3 and S4). The most common pathogen detected was diarrheagenic *E. coli* (248 organisms detected among 187 patients) followed by norovirus (144 patients) and *Campylobacter* (114 patients) (Table 3).

**Table 2.**
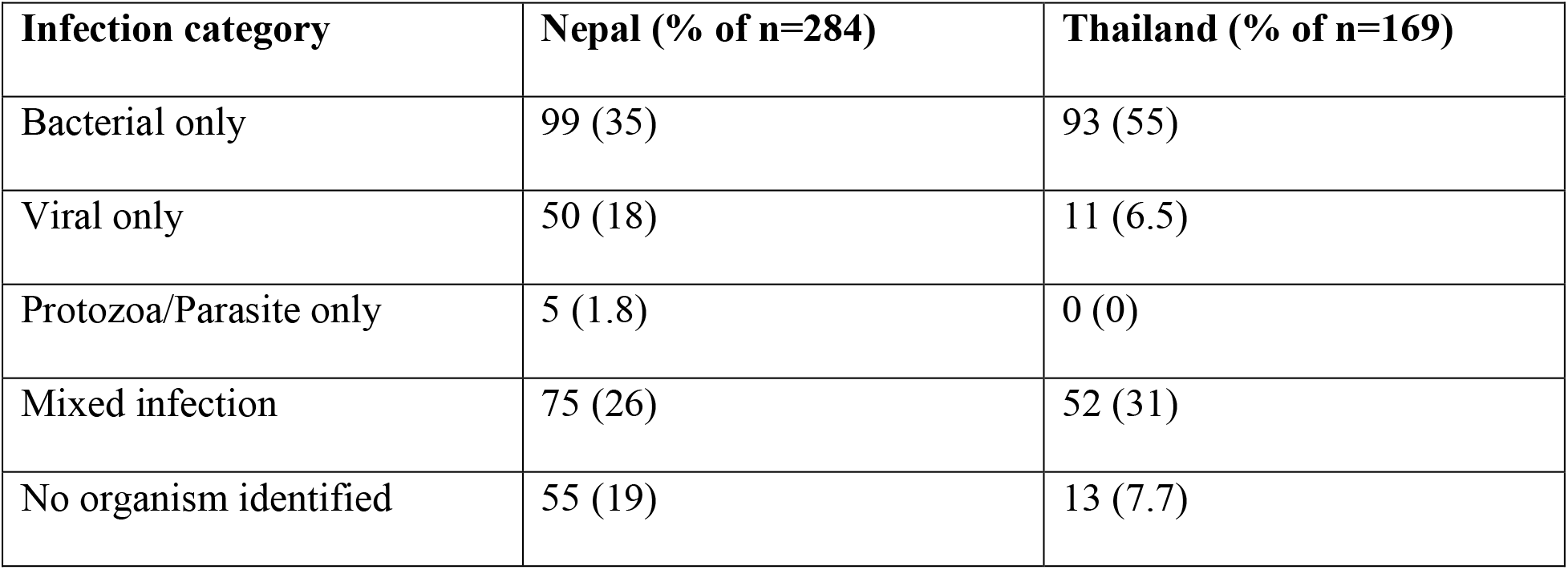
Stool analysis results showing categories of infection types and number of infections per site in each category.

| **Infection category** | **Nepal (% of n=284)** | **Thailand (% of n=169)** |
| --- | --- | --- |
| Bacterial only | 99 (35) | 93 (55) |
| Viral only | 50 (18) | 11 (6.5) |
| Protozoa/Parasite only | 5 (1.8) | 0 (0) |
| Mixed infection | 75 (26) | 52 (31) |
| No organism identified | 55 (19) | 13 (7.7) |

**Table 3.** Number of detected pathogens among all patient cases.

| Pathogen | Number detected |
| --- | --- |
| Diarrheagenic *E. coli* | 248 |
| ETEC | 85 |
| EAEC | 73 |
| EPEC | 72 |
| Misc. E coli (EIEC, or<br>STEC) | 18 |
| Norovirus | 144 |
| *Campylobacter* | 114 |
| No pathogens | 68 |
| Sapovirus | 47 |
| *Aeromonas* | 42 |
| *Plesiomonas* | 40 |
| *Salmonella* | 36 |
| *Shigella* | 34 |
| Rotavirus | 15 |
| *Vibrio* | 13 |
| *Bacteroides* | 10 |
| Miscellaneous parasitic | 14 |
| Miscellaneous bacteria | 10 |
| Miscellaneous viruses | 7 |

We analyzed multiple model variations to identify predictors of bacterial TD. Using the same variable selection process, variations comparing bacterial-only etiologies to viral plus non-detected infections outperformed models using total bacterial cases (bacterial-only infections plus mixed infections with bacterial and non-bacterial pathogens) in cross-validation. Given our goal of identifying the strongest predictors of bacterial diarrhea as well as the challenge of identifying the predominant pathogen among mixed infections, we subsequently excluded cases of mixed infections from analysis.

Using random forest regression, we identified predictors of bacterial diarrhea and ranked them from strongest to weakest predictors based off of their reduction in mean squared prediction error (Table 4). Average environmental temperature (Odds ratio (OR) 0.99, 95% Confidence Interval (CI) 0.98-1.0), and RBC on stool microscopy (OR 1.27, CI 1.13-1.43) were the top predictors of diarrheal etiology. When separated by site, using univariable analysis, we found that the variables with the most evidence of an association with bacterial etiology at the Nepal site included RBCs on stool microscopy, absence of fever, and a lower average site temperature (Table S2), and at the Thai site included the presence of mucus on stool microscopy, an absence of vomiting, and the presence of RBC on stool microscopy (Table S3).

**Table 4.** The top 10 predictive variables based on random forest ranked in descending order based on mean squared prediction error (IncMSE). Each variable is shown with its associated IncMSE, odds ratio, and P value.

|  | **Variable** | **Mean Square-Error**<br>**(IncMSE)** | **Odds ratio** | **P value** |
| --- | --- | --- | --- | --- |
| **1** | Average environmental temperature | 22.39 | 0.99 (0.98 – 1.00) | 0.001 |
| **2** | Stool study: RBC | 16.19 | 1.27 (1.13 - 1.43) | <0.001 |
| **3** | Total # of diarrheal stools at time of evaluation | 6.66 | 1 (0.99 – 1.00) | 0.11 |
| **4** | Nationality: North American | 3.87 | 0.95 (0.83 - 1.10) | 0.505 |
| **5** | Age | 3.24 | 1.01 (1.00 - 1.01) | 0.009 |
| **6** | Average environmental precipitation | 2.64 | 1.18 (0.80 - 1.74) | 0.406 |
| **7** | Nationality: European | 1.91 | 1.12 (0.99 - 1.27) | 0.078 |
| **8** | Stool study: WBC | 1.84 | 1.06 (0.91 - 1.23) | 0.444 |
| **9** | Stool grade: loose | 1.45 | 1.04 (0.94 - 1.16) | 0.448 |
| **10** | Stool study: mucus | 0.41 | 1.02 (0.90 - 1.14) | 0.783 |

In order to identify models that would balance a high AUC with ease of use, we selected the three model variations below. We started with clinical and demographic data and initially excluded weather and stool microscopy data as these data may be challenging for travelers to obtain. Weather data was included in our second model iteration and stool microscopy included in the third given that stool studies represent the most challenging variables for a traveler to obtain. Our final three model variations included:

1. Model 1: Patient symptoms and demographic variables only, no weather or stool microscopy variables
2. Model 2: Symptoms, demographics, and weather variables without stool microscopy
3. Model 3: All variables including weather and stool microscopy.

In 5-fold cross-validation, we found that the inclusion of weather and microscopy variables yielded an increase in the maximum discriminative ability among all models. Maximum discriminative ability with only symptoms and history (Model 1) occurred at 8 variables with an AUC of 0.0.63 using LR (calibration intercept -0.01 (SD 0.30) and slope 0.57 (SD 0.44), Figure 1A). Maximum discriminative ability with inclusion of weather data (Model 2) occurred at 1 variable with an AUC of 0.70 using LR (calibration intercept -0.05 (SD 0.30) and slope 1.21 (SD0.65), Figure 1B). The inclusion of both weather and stool microscopy variables (Model 3) demonstrated an AUC of 0.73 at 3 variables using LR (calibration intercept -0.01 (SD 0.31) and slope 0.95 (SD 0.36), Figure 1C). Overall, Model 3 demonstrated the highest AUC and best calibration, with logistic methods demonstrating a higher AUC compared to RF methods. In sensitivity analysis, a prediction model discriminating Shigella/Campylobacter infections from non-bacterial infections resulted in a maximum AUC of 0.75 (Figure S4).

**Figure 1.**
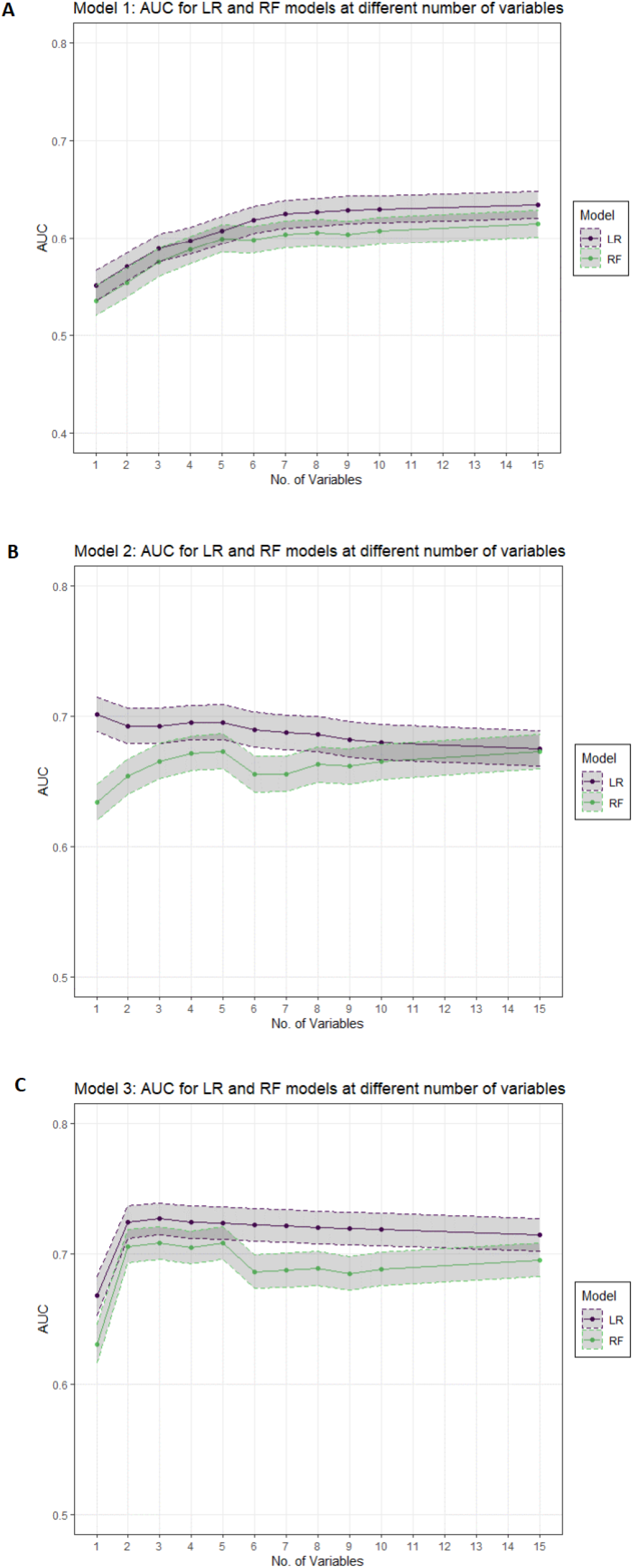
Area under the ROC curve (AUC) for bacterial etiology obtained at a range of variables using logistic regression (LR) and random forest (RF) methods. A) Model 1 includes demographic and symptom variables and excludes weather and stool microscopy variables. Maximum predictive accuracy in Model 1 occurred at 9 variables using LR (AUC 0.70, calibration intercept 0.2 (SD 0.32) and slope 0.74 (SD 0.39)). B) Model 2 includes demographic, symptom, and weather variables while excluding stool microscopy variables. Maximum predictive accuracy in Model 2 occurred at 2 variables using LR (AUC 0.71, calibration intercept 0.04 (SD 0.29) and slope 1.05 (SD 0.57)). C) Model 3 includes all demographic, symptom, weather, and stool microscopy variables. Maximum predictive accuracy in Model 3 occurred at 3 variables using LR (AUC 0.74, calibration intercept 0 (SD 0.31) and slope 1 (SD 0.36)).

## Discussion

Diarrhea is the most common health complaint among international travelers, and one that occurs more frequently in travelers to low and middle-income countries (LMICs)^1, 20^. Current guidelines recommend traveler-initiated self-treatment of moderate to severe diarrhea, with limited consideration of etiology; thus, tools differentiating bacterial from non-bacterial etiologies can provide practical support for antibiotic use decision-making. In this study, we used data from two studies of traveler’s diarrhea to derive clinical predictors of bacterial etiology.

Our model identified the top predictors of bacterial diarrhea to be a lower location-specific environmental temperature and the presence of red blood cells (RBCs) on stool microscopy. A recent study of 1450 patients evaluated for TD in the UK found male gender, younger age, and WBC on microscopy to be predictive of bacterial disease ^20^, while a non-traveler-specific study found associations of bacterial diarrhea with bloody stools, fevers, lack of vomiting, and longer diarrheal duration ^21^. Neither of these studies considered weather variables among potential predictors, and our demonstration of the importance of a location-specific variable such as environmental temperature, suggests that this should be included in future predictive modeling efforts.

We found that a 14-day average of environmental temperature and precipitation increased model performance. While prior studies have documented complex seasonal patterns among diarrheal diseases in tropical climates ^2, 8, 22^, studies examining weather data for individual-level clinical prediction are lacking. Climate is thought to aid pathogen transmission through mechanisms such as contamination of food or water supplies, pathogen survival on fomite surfaces, or facilitating vector life cycles. Among data collected for the Global Enteric Multicenter Study (GEMS), bacterial pathogens demonstrated higher prevalence during the hot and/or rainy seasons ^2^ while viral etiologies demonstrated higher prevalence during the dry/cold season ^8, 12^. For translation of prediction models into decision support tools, weather variables could be gathered from online weather sources, based on smartphone-based detection of GPS location.

We showed that top predictors differed by site, which is not surprising and may suggest a need for location-specific data in generating clinical prediction models. Prior analyses of data from the GeoSentinel travel clinics demonstrate the significant role of travel location on the risk of TD development^23, 24^. Overall, models including weather variables (Model 2) or both weather and stool microscopy variables (Model 3) demonstrated higher discriminatory performance based on cross-validation compared to models with only symptomatic and demographic variables (Model 1). When included, RBCs on stool microscopy appear as the 2nd most important predictive variable. Blood in stool is known to be a specific yet insensitive predictor of bacterial diarrhea ^25-27^. Unfortunately, not all travelers have access to stool microscopy testing, though the use of a point of care stool RBC diagnostic such as used for colon cancer screening deserves further consideration. While stool RBCs may improve model accuracy, it is unclear whether the gain in performance (increase in AUC of 0.03) is worth the resources needed for stool microscopy.

Our study has several limitations. First, we acknowledge that our inclusion of only two hospital sites in Asia limit interpretation and generalizability. Exclusion of mixed-organism infections further reduces generalizability and led to the exclusion of 127 cases. Secondly, not all bacterial pathogens included in analysis would necessarily require antibiotics, though studies are lacking regarding the etiologies that would benefit most. While infection with *Salmonella, Shigella* and *Campylobacter spp*. are thought to benefit from antibiotics ^28^, there is also evidence from randomized controlled trials in travelers that treatment of diarrheagenic *E. coli* reduces symptom duration by up to 2 days. Even among diarrheagenic *E. coli*, certain strains such as EAEC or EPEC may be less clinically meaningful. Future models may further differentiate between treatable and non-treatable pathogens, particularly following improvements in diagnostic testing. Third, our use of a set cycle threshold (<35) for determination of causative pathogen may be overly sensitive and contributed to a higher number of mixed-organism infections. Lastly, our results were only cross-validated and future work is required to externally validate and implement a clinical prediction rule based on the predictors identified in this study.

Nevertheless, we demonstrate that location-specific weather variables influence diarrheal etiology and should be considered in future work on clinical prediction and decision support tools for travelers’ diarrhea.

## Data Availability

All data produced in the present study are available upon reasonable request to the authors.

## Acknowledgements

This work was supported in part by the NIAID of the NIH under award number R01AI135114 (to D.T.L.), and by the University of Utah Study Design and Biostatistics Center, with funding in part from the National Center for Research Resources and the National Center for Advancing Translational Sciences, National Institutes of Health, through Grant UL1TR002538 (formerly 5UL1TR001067-05, 8UL1TR000105 and UL1RR025764).

Material has been reviewed by the Walter Reed Army Institute of Research (WRAIR). There is no objection to its presentation and/or publication. The opinions or assertions contained herein are the private views of the author, and are not to be construed as official, or as reflecting true views of the Department of the Army or the Department of Defense. The investigators have adhered to the policies for protection of human subjects as prescribed in AR 70–25.

## Author Contributions

Study Design: LB, DTL. Data Collection: PP, SShrestha, SA, SD, SSornsakrin, JAP, LB. Analysis: MAP, TS, BJB, JAP, DTL. Interpretation: MAP, TS, BJB, PP, SShrestha, SA, SD, SSornsakrin, JAP, LB, DTL.

